# Distinct clinical and biological characteristics of acute myeloid leukemia with higher expression of long noncoding RNA *KIAA0125*

**DOI:** 10.1101/2020.08.21.20179267

**Authors:** Yu-Hung Wang, Chien-Chin Lin, Chia-Lang Hsu, Sheng-Yu Hung, Chi-Yuan Yao, Sze-Hwei Lee, Cheng-Hong Tsai, Hsin-An Hou, Wen-Chien Chou, Hwei-Fang Tien

## Abstract

Expression of long non-coding RNA *KIAA0125* has been incorporated in various gene expression signatures for prognostic prediction in acute myeloid leukemia (AML) patients, yet its functions and clinical significance remain unclear. This study was aimed to investigate the clinical and biological characteristics of AML bearing different levels of *KIAA0125*. We profiled *KIAA0125* expression levels in bone marrow cells from 347 *de novo* AML patients and found higher *KIAA0125* expression was closely associated with *FLT3-ITD, RUNX1*, and *DNMT3A* mutations, and M1 subtype by the French-American-British classification, but inversely correlated with t(8;21) and t(15;17). Among the 227 patients who received standard chemotherapy, those with higher *KIAA0125* expression had a lower complete remission rate, shorter overall survival (OS) and disease-free survival (DFS) than those with lower expression. The prognostic significance was validated in both TCGA and GSE12417 cohorts. Subgroup analyses showed that higher *KIAA0125* expression also predicted shorter DFS and OS in patients with normal karyotype or non-M3 AML. In multivariable analysis, higher *KIAA0125* expression remained an adverse risk factor independent of age, WBC counts, karyotypes, and mutation patterns. Bioinformatics analyses revealed that higher *KIAA0125* expression was associated with hematopoietic and leukemic stem cell signatures and ATP-binding cassette transporters, two predisposing factors for chemoresistance.

## Introduction

Long non-coding RNAs (lncRNAs) are non-protein coding RNAs that are longer than 200 nucleotides. Comparing to other classes of ncRNAs, lncRNAs exhibit a wide range of structures and functions[1]. Recently, lncRNAs have emerged as important regulators for gene expression via remodeling nuclear architecture, modulating mRNA stability and translation, and post-translational modifications[1–4]. Besides, some lncRNAs are dysregulated and harbor prognostic relevance in several types of cancers[5–8]. However, the roles of lncRNAs in tumorigenesis are still largely unknown.

In recent years, several gene expression-based prognostic scores have been developed for better risk stratification of acute myeloid leukemia (AML) patients[9–14]. Among those high-risk genes, lncRNA gene *KIAA0125* (also named as *FAM30A)* is unique because it is the only non-coding gene and is expressed in humans but not in mice (From the UniProt database, https://www.uniprot.org/uniprot/Q9NZY2). Additionally, *KIAA0125* expression was integrated into a recently proposed 17-gene stemness score, which could predict outcomes in AML patients[9].

In this study, we aimed to investigate the association of *KIAA0125* expression with clinical and biological characteristics in AML patients. We first profiled the expression levels of *KIAA0125* in bone marrow (BM) cells from AML patients and normal controls and demonstrated that AML patients had higher *KIAA0125* expression than normal controls. Higher expression of *KIAA0125* was associated with distinct clinical and biological characteristics and served as an independent poor prognostic biomarker for AML patients in ours and two other publicly annotated cohorts. Further bioinformatics analyses showed that higher expression of *KIAA0125* in AML was closely associated with hematopoietic stem cell (HSC) and leukemic stem cell (LSC) signatures and several important ATP-binding cassette transporters (ABC transporters); these factors are regarded responsible for chemoresistance in AML. *KIAA0125* could be a potential target for novel treatment in AML patients with high *KIAA0125* expression.

## Materials and Methods

### Patients

We recruited 347 adult patients with *de novo* AML diagnosed in the National Taiwan University Hospital (NTUH) from 1996 to 2011 who had enough cryopreserved BM cells for tests. The diagnoses were based on the French-American-British (FAB) and the 2016 World Health Organization classifications[15,16]. Among them, 227 patients received standard chemotherapy. Non M3 (acute promyelocytic leukemia, APL) patients received idarubicin 12 mg/m^2^ per day days 1–3 and cytarabine 100 mg/m^2^ per day days 1–7, and then consolidation chemotherapy with 2–4 courses of high-dose cytarabine 2000 mg/m^2^ q12h for total 8 doses, with or without an anthracycline (Idarubicin or Mitoxantrone), after achieving complete remission (CR) as described previously[17]. APL patients received concurrent all-trans retinoic acid and chemotherapy. The remaining 120 patients received supportive care and/or reduced-intensity anti-leukemia therapy due to underlying comorbidities or based on the decision of the physicians or patients. BM samples from 30 healthy donors of hematopoietic stem cell transplantation (HSCT) were collected as normal controls. This study was approved by the Institutional Review Board of the NTUH.

## Microarray and genetic alteration analysis

We profiled the global gene expression of BM mononuclear cells from 347 AML patients and 30 healthy transplant donors by Affymetrix GeneChip Human Transcriptome Array 2.0 as described previously[11, 18, 19]. The raw and normalized microarray data reported in this article have been deposited in the Gene Expression Omnibus database (accession number GSE68469 and GSE71014)[11,18,19]. For external validation, we analyzed microarray datasets of GSE12662 (n = 76) and GSE12417-GPL96 cohorts (n = 163), and RNAseq dataset of the TCGA cohort (n = 186)[20–22]. Cytogenetic analyses were performed and interpreted as described previously[23]. We also analyzed the mutation statuses of 17 myeloid-relevant genes including *ASXL1, IDH1, IDH2, TET2, DNMT3A, FLT3-ITD, FLT3-TKD, KIT, NRAS, KRAS, RUNX1, MLL/PTD, CEBPA, NPM1, PTPN11, TP53*, and *WT1* by Sanger sequencing as previously described[17, 18, 23–26].

## Analysis of gene expression in next-generation sequencing datasets

We analyzed gene expression data of 141 AML samples profiled with Illumina Genome Analyzer RNA Sequencing in the TCGA database[22] to investigate the absolute gene expression levels.

## Gene Set Enrichment Analysis

The preranked Gene Set Enrichment Analysis (GSEA) implemented by R package clusterProfiler was performed using the stem cell-related gene sets from the MSigDB databases. The genes were ranked based on the Spearman’s correlation coefficient between the given gene and *KIAA0125*.

## Statistical analysis

We used the Mann-Whitney U test to compare continuous variables and medians of distributions. The Fisher exact test or the x2 test were performed to examine the difference in discrete variables, including gender, FAB classification, cytogenetic changes, and genetic alterations between patients with lower and higher *KIAA0125* expression. Overall survival (OS) was the duration from the date of initial diagnosis to the time of last follow-up or death from any cause, whichever occurred first. Disease free survival (DFS) was the duration from the date of attaining leukemia-free state until the date of AML relapse or death from any cause, whichever occurred first. The survival prediction power of *KIAA0125* expression was evaluated by both the log-rank test and the univariate Cox proportional hazards model. We plotted the survival curves with Kaplan-Meier analysis and calculated the statistical significance with the log-rank test. The Cox proportional hazards model was used in multivariable regression analysis. P values < 0.05 were considered statistically significant. All statistical analyses were performed with BRB-ArrayTools (version 4.5.1; Biometric Research Branch, National Cancer Institute, Rockville, MD), and IBM SPSS Statistics 23 for Windows.

## Results

The median age of the 347 AML patients was 57 years. Among the 331 patients who had cytogenetic data at diagnosis, 165 (49.8%) had clonal chromosomal abnormalities. Sixty patients (18.1%) had favorable cytogenetics; 223 (67.2%), intermediate-risk cytogenetics; and 14.8%, unfavorable cytogenetics (Supplement Table 1) based on the refined British Medical Research Council (MRC) classification[27]. The clinical and laboratory characteristics of these patients at diagnosis are summarized in Table 1.

**Table 1.** Comparison of clinical and laboratory features between AML patients with lower and higher BM *KIAA0125* expression

| Clinical characters | Total (N=347) | High *KIAA0125* (n=174) | Low *KIAA0125* (n=173) | P value |
| --- | --- | --- | --- | --- |
| **Sex** |  |  |  | 0.174 |
| Male | 196 | 92 | 104 |  |
| Female | 151 | 82 | 69 |  |
| **Age\*** |  | 57 (15-91) | 58 (18-90) | 0.830 |
| **Laboratory data\*** |  |  |  |  |
| WBC, X 10<sup>9</sup> /L | 21.9 (0.38-423) | 21.4 (0.38-417.5) | 22.38 (0.65-423.0) | 0.872 |
| Hb, g/dL | 8.1 (3.3-16.2) | 8.1 (3.3-13.2) | 8.1 (3.7-16.2) | 0.959 |
| Platelet, X 10<sup>9</sup> /L | 45 (2-655) | 54 (6-455) | 41 (2-655) | 0.060 |
| Blast, X 10<sup>9</sup> /L | 9.1 (0-369.1) | 12.3 (0-345.9) | 5.7 (0-369.1) | 0.021 |
| LDH (U/L) | 917 (202-13130) | 892.5 (242-7734) | 925 (202-13130) | 0.787 |
| **FAB classification, n (%)** |  |  |  |  |
| M0 | 6 | 4 (2.4) | 2 (1.2) | 0.414 |
| M1 | 67 | 48 (28.4) | 19 (11.1) | <0.001 |
| M2 | 109 | 56 (33.1) | 53 (30.8) | 0.756 |
| M3 | 28 | 3 (1.8) | 25 (14.5) | <0.001 |
| M4 | 103 | 49 (28.9) | 54 (31.4) | 0.534 |
| M5 | 20 | 6 (3.6) | 14 (8.1) | 0.063 |
| M6 | 8 | 3 (1.8) | 5 (2.9) | 0.469 |
| Undetermined | 6 | 5 | 1 | 0.215 |
| **Induction response, n (%)** | 227 | 116 | 111 |  |
| CR | 165 (72.7) | 71 (61.2) | 94 (84.7) | <0.001 |
| PR | 5 (2.2) | 4 (3.4) | 1 (0.9) | 0.191 |
| Refractory | 42 (18.5) | 33 (28.4) | 9 (8.1) | <0.001 |
| Induction death | 15 (6.6) | 8 (6.9) | 7 (6.3) | 0.858 |
| Relapse (%) | 72 (31.7) | 42 (36.2) | 30 (27.0) | 0.137 |

Abbreviations: CR, complete remission; Hb, hemoglobin; HSCT, allogeneic hematopoietic stem cell transplantation; LDH, lactate dehydrogenase; PR, partial remission.
\*Median (range).

## Comparison of clinical characteristics and genetic alterations between patients with higher and lower *KIAA0125* expression

We first compared the BM *KIAA0125* expression between the 30 healthy controls and 347 AML patients. The expression of *KIAA0125* was significantly higher in AML samples than healthy controls (*p*< 0.001, Figure 1a). Then, the 347 AML patients were divided into two groups by the median value of the *KIAA0125* expression. The comparison of clinical and laboratory features between the two groups is shown in Table 1. The higher *-KIAA01Z25* group had higher circulating blasts at diagnosis (*p* = 0.021) and higher incidence of FAB M1 subtype (*p*< 0.001), but lower incidence of M3 subtype (*p*< 0.001), compared to the lower*-KIAA0125* group (Table 1). Patients with FAB immature subtype of M0 or M1 had higher *KIAA0125* expression while those with M3 subtype had lower expression of *KIAA0125* (*p*< 0.001, Figure 1c), and similar results were noted in the GSE12662 cohort (*p*< 0.001, Figure 1d). Moreover, higher*-KIAA0125* patients had significantly lower frequencies of t(8;21) and t(15;17) in both NTUH cohort,(both *p*< 0.001, Supplement Table 1) and TCGA cohort (*p*= 0.006) and (*p*< 0.001, respectively, Supplement Table 1). The higher*-KIAA0125* patients more frequently had *FLT3-ITD* (*p* = 0.048) and mutations in *DNMT3A* (*p* = 0.015), and *RUNX1* (*p*= 0.034) (Supplement Table 2). Compatible with this finding, patients with *DNMT3A* or *RUNX1* mutation had higher *KIAA0125* expression than those without the mutation *(p* = 0.019 and 0.045, respectively, Supplement Figure 1). Similarly, there was close association between higher *KIAA0125* expression and *DNMT3A* (*p*= 0.001) and *RUNX1* mutations (*p*= 0.017) in the TCGA cohort (Supplement Table 3). Among the 227 patients who received standard chemotherapy, 166 (73.1%) patients attained a complete remission (CR) while 43 (18.9%) patients had primary refractory diseases. Notably, the patients with higher *KIAA0125* expression had a lower CR rate (61.2% vs. 84.7%, *p*< 0.001) than those with lower expression. In accordance with this finding, the patients who achieved CR after induction chemotherapy had lower expression of BM *KIAA0125* at diagnosis than those who did not (*p*< 0.001, Figure 1b).

**Figure 1.**
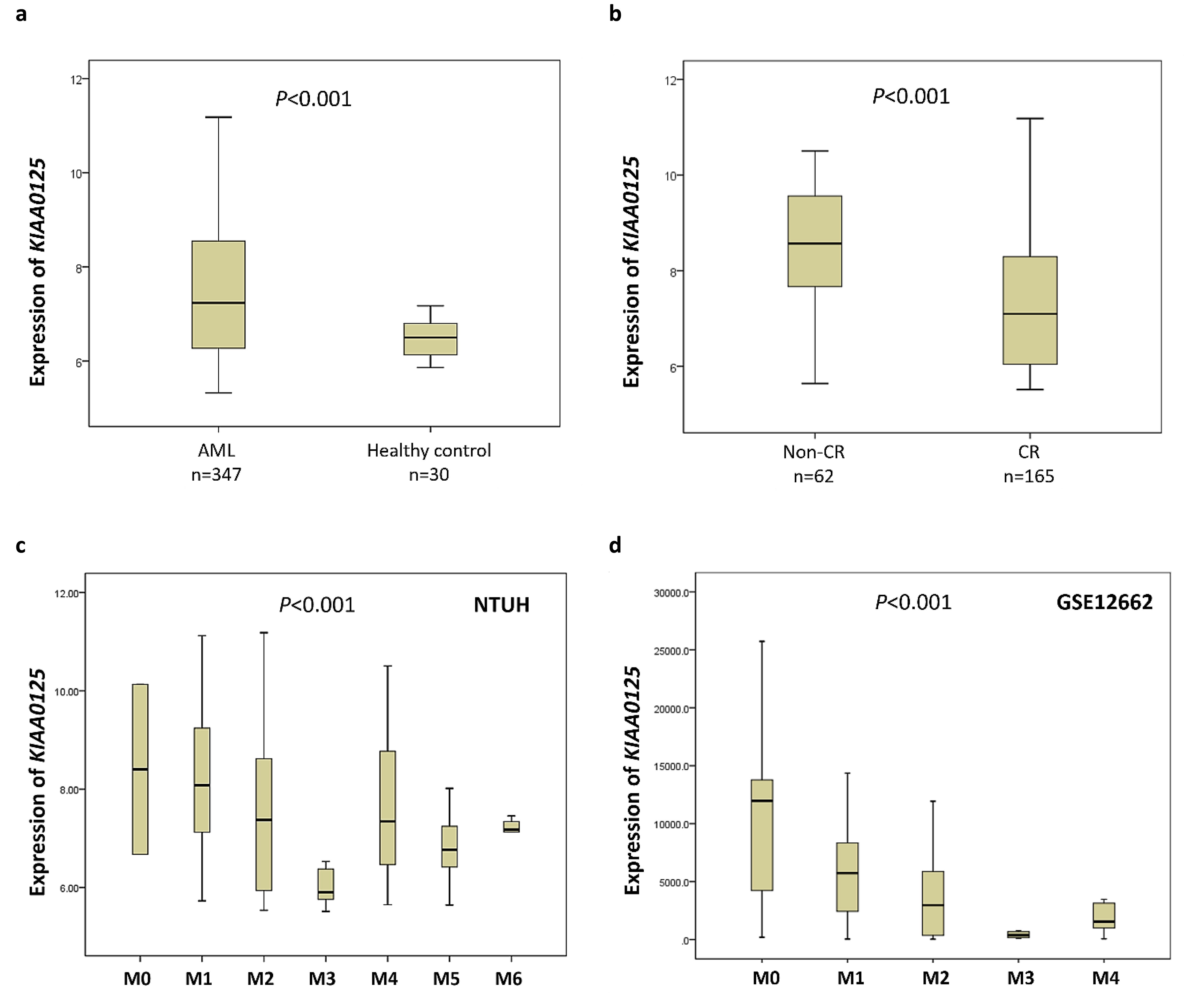
Box plots depicting expression levels of KIAA0125 in healthy controls and various AML subgroups. (a) Patients with AML had significantly higher expression of *KIAA0125* than healthy controls; (b) patients who achieved CR after induction chemotherapy had lower expression of BM *KIAA0125* at diagnosis than those who did not; and (c, d) patients with AML, M3 had significantly lower expression of *KIAA0125* while those with more immature subtype M0 or M1 had significantly higher expression in both NTUH cohort (c) and GSE12662 cohort (d).

## The impacts of the *KIAA0125* expression on OS and DFS

Patients with higher *KIAA0125* expression had an inferior DFS and OS than those with lower expression, no matter whether the survival was censored on the day of hematopoietic stem cell transplantation (HSCT) (median, 11.7 months vs. 101.7 months, (*p*< 0.001; and 20 months vs not reached (NR), (*p* = 0.001, respectively; Figure 2a and 2b) or not (*p* = 0.001 and *p* = 0.001, respectively; Supplement Figure 2a and 2b). Subgroup analyses showed that the prognostic significance of *KIAA0125* expression for DFS and OS remained valid in both non-APL and normal karyotype patients (Figure 2 and 2d).

**Figure 2.**
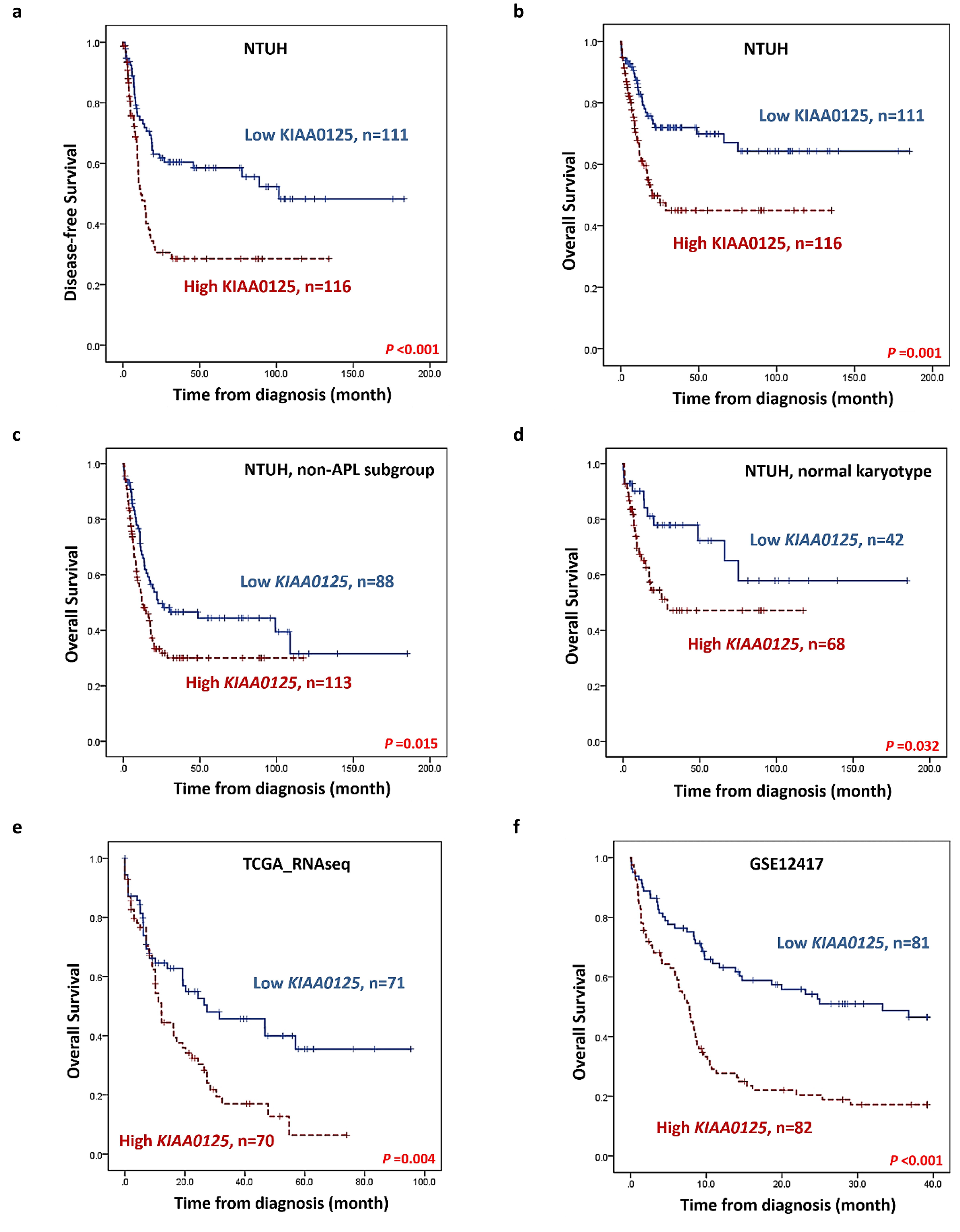
Kaplan-Meier survival curves stratified by expression of *KIAA0125*. DFS (a) and OS (b) of the 227 AML patients receiving standard chemotherapy in the NTUH cohort; OS of 201 non-APL patients (c) and 110 cytogenetically normal AML patients (d) who received standard treatment in the NTUH cohort; and OS of 141 patients in the TCGA cohort (e) and GSE12417-GPL96 cohort. Patients with higher *KIAA0125* expression had worse clinical outcomes than those with lower expression.

In multivariable analysis, we included clinically relevant parameters and variables with a (*p*-value < 0.05 in univariate Cox regression analysis (Supplement Table 4) as covariates, including age, white blood cell counts at diagnosis, karyotypes, mutation statuses of *NPM1/FLT3-ITD, CEBPA^double^^mutations^_;_ RUNX1, MLL-PTD*, and *TP53*, and *KIAA0125* expression. Higher *KIAA0125* expression, either divided by a median (Table 2) or calculated as continuous values (Supplement Table 5), was an independent adverse prognostic factor for DFS (*p*< 0.001) and (*p*< 0.001, respectively) and OS (*p* = 0.002 and (*p* = 0.001, respectively). To verify the prognostication power of the *KIAA0125* expression, we analyzed the expression of *KIAA0125* and its prognostic significance in the TCGA cohort and the GSE12417-GPL96 cohort. Consistent with the findings in the NTUH cohort, patients with higher *KIAA0125* expressions had a significantly shorter OS (12.2 months vs. 27.4 months, (*p* = 0.004, and 7.8 months vs. 33.3 months, (*p*< 0.001, respectively, Figure 2e and 2f) than those with lower *KIAA0125* expression in the two external validation cohorts.

**Table 2.** Multivariable analysis for DFS and OS in 227 AML patients who received standard intensive chemotherapy

| Variable | DFS |  |  |  | OS |  |  |  |
| --- | --- | --- | --- | --- | --- | --- | --- | --- |
|  | 95% CI |  |  |  | 95% CI |  |  |  |
|  | HR | Lower | Upper | P | HR | Lower | Upper | P |
| Age* | 1.009 | 0.997 | 1.021 | 0.129 | 1.031 | 1.014 | 1.047 | <0.001 |
| WBC* | 1.004 | 1.001 | 1.007 | 0.002 | 1.005 | 1.001 | 1.008 | 0.009 |
| Karyotype† | 1.762 | 1.310 | 2.369 | <0.001 | 1.910 | 1.288 | 2.833 | 0.001 |
| *NPM1*/*FLT3*-ITD‡ | 0.540 | 0.297 | 0.981 | 0.043 | 0.812 | 0.404 | 1.634 | 0.559 |
| *CEBPA*<sup>double</sup> | 0.423 | 0.216 | 0.830 | 0.012 | 0.253 | 0.077 | 0.833 | 0.024 |
| *RUNX1* | 1.463 | 0.841 | 2.546 | 0.178 | 1.492 | 0.763 | 2.914 | 0.242 |
| *MLL*-PTD | 2.864 | 1.346 | 6.094 | 0.006 | 3.017 | 1.136 | 8.017 | 0.027 |
| *TP53* | 1.691 | 0.607 | 4.705 | 0.315 | 2.798 | 0.875 | 8.950 | 0.083 |
| Higher *KIAA0125* expression§ | 2.609 | 1.760 | 3.867 | <0.001 | 2.226 | 1.335 | 3.711 | 0.002 |

Abbreviations: HR, hazard ratios; CI, confidence interval.
\*As continuous variable.
†Unfavorable cytogenetics versus others. The classification of favorable, intermediate and unfavorable
cytogenetics is based on the refined Medical Research Council (MRC) classification [27]. Favorable:
t(15;17)(q22;q21), t(8;21)(q22;q22), and inv(16)(p13q22)/t(16;16)(p13;q22); unfavorable: abn(3q) (excluding
t(3;5)(q25;q34)), inv(3)(q21q26)/t(3;3)(q21;q26), add(5q)/del(5q), -5, -7, add(7q)/del(7q), t(6;11)(q27;q23),
t(10;11)(p11;q23), other t(11q23) (excluding t(9;11)(p21~22;q23) and t(11;19)(q23;p13)), t(9;22)(q34;q11),
-17, and abn(17p); and intermediate: entities not classified as favorable or adverse. Seven patients without
chromosome data were not included in the analysis.
‡*NPM1*+/*FLT3*-ITD- versus other subtypes.
§High vs. low expression of *KIAA0125* (median as cutoff)

## Biological impacts of *KIAA0125* in AML

To gain biological insights into the underlying mechanism of unfavorable prognosis related to *KIAA0125* overexpression, we investigated the genes whose expression are strongly correlated with that of *KIAA0125*. Since *KIAA0125* was reported as an LSC marker, we curated several published HSC and LSC signatures from different studies[28–30]. GSEA showed HSC and LSC signatures were all significantly enriched in the patients with higher *KIAA0125* expression in both the NTUH and TCGA cohorts (both *(p*< 0.001, Figure 3a). We next checked the leading-edge genes whose expression levels were most positively correlated to *KIAA0125* expression in both NTUH and TCGA cohort. Among them, *SPINK2, MAP7, HOPX, MMRN1, DNMT3B, TCF4, SLC38A1, DOCK1, ARHGAP22, MN1*, and 4 genes in the ATP-binding cassette (ABC) superfamily *(ABCG1, ABCA2, ABCB1*, and *ABCC1)* have been reported to be associated with poor prognosis or chemoresistance in AML (Figure 3b and Table 3)[9, 31–50].

**Figure 3.**
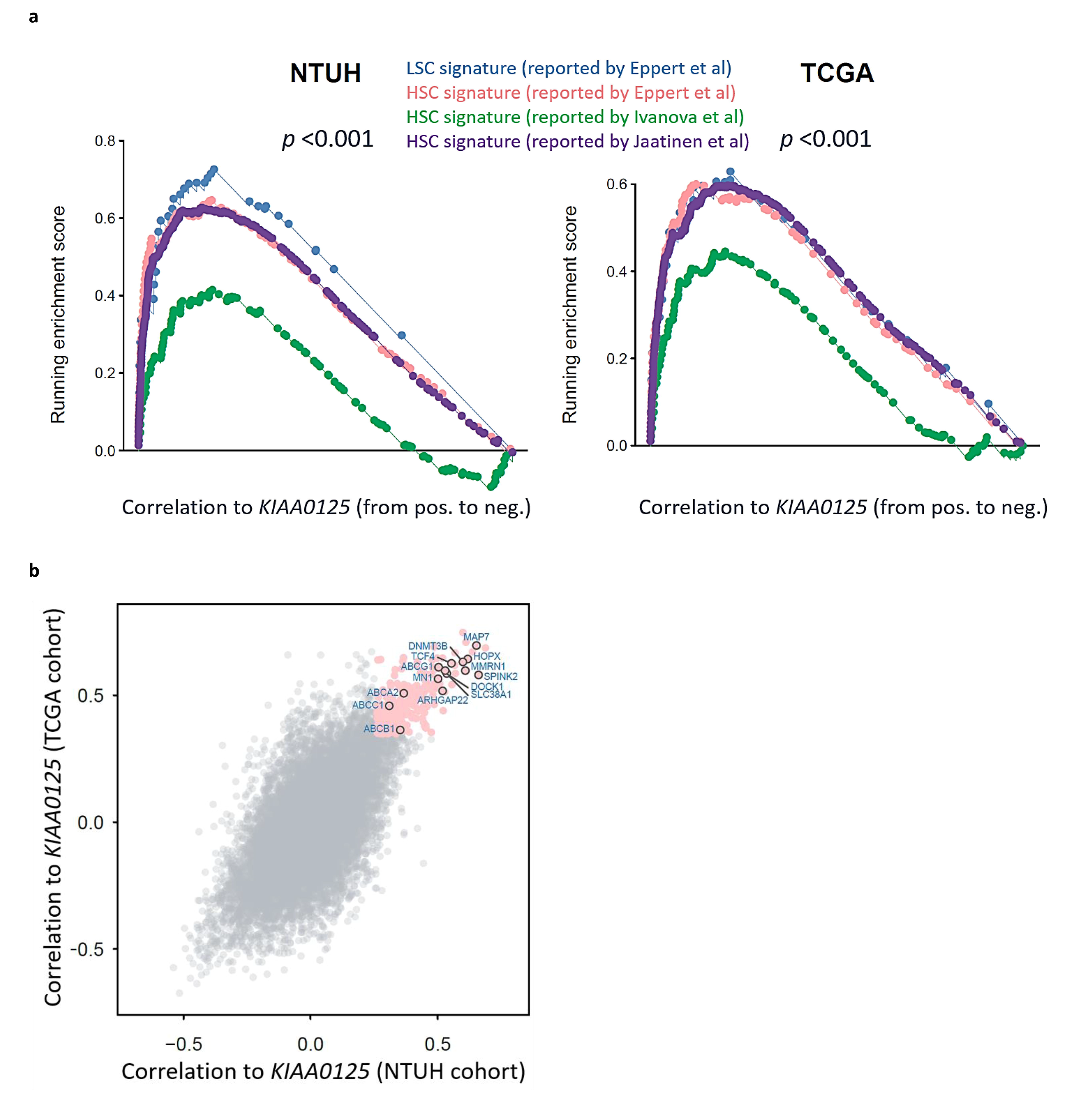
GSEA enrichment plots of HSC and LSC signatures and scatter plot of genes positively associated with higher *KIAA0125* expression. (a) GSEA enrichment plots show positive association of higher *KIAA0125* expression with HSC and LSC signatures curated from several published reports in both the NTUH and TCGA cohorts; (b) the scatter plot reveals the genes strongly correlated to *KIAA0125* expression in both the NTUH and TCGA cohorts (pink). The correlation measurement is based on the Spearman’s correlation coefficient between the given gene and *KIAA0125*. The strongly correlated genes are defined as their correlation values at top 5% of all genes in both cohorts.

**Table 3.** Summary of the biological functions of the *KIAA0125*-associated genes that have been reported to be associated with prognosis or drug resistance in AML patients and their correlation values with *KIAA0125* in ours and the TCGA cohort

| Genes | Correlation coefficient (*p* value) |  |  |  | Association with leukemia |
| --- | --- | --- | --- | --- | --- |
|  | NTUH |  | TCGA |  |  |
| *SPINK2* | 0.661 | (3.4E-45) | 0.5798 | (6.2E-15) | Serine Peptidase Inhibitor; upregulation is associated with poor outcomes in adult patients with AML[31]; integrated into a 6-gene LSC score to identifies high risk pediatric AML[32] |
| *MAP7* | 0.653 | 1.0E-43 | 0.696 | (<E-45) | Microtubule-associated proteins, overexpressed in cytogenetically normal AML patients with dismal outcomes[33] |
| *HOPX* | 0.619 | (2.6E-38) | 0.643 | (<E-45) | The smallest homeodomain protein; higher expression predicts poor prognosis in *de novo* AML[34] |
| *MMRN1* | 0.609 | (9.7E-37) | 0.597 | (<E-45) | A member of the elastin microfibrillar interface protein; an adverse marker in both pediatric and adult AML[35] |
| *DNMT3B* | 0.599 | (1.7E-35) | 0.631 | (<E-45) | DNA methyltransferases; an important LSC marker[36-38] |
| *TCF4* | 0.556 | (1.1E-29) | 0.626 | (<E-45) | A transcription factor; predict outcome in *RUNX1* mutated and translocated AML[39,40] |
| *SLC38A1* | 0.536 | (2.3E-27) | 0.585 | (<E-45) | A glutamine amino acid transporter, overexpressed in AML patients with adverse clinical outcomes[41] |
| *DOCK1* | 0.530 | (1.1E-26) | 0.597 | (5.9E-16) | A novel class of guanine nucleotide exchange factors; high expression confers poor prognosis in AML[42] |
| *ARHGAP22* | 0.519 | (1.5E-25) | 0.518 | (<E-45) | Rho GTPase activating protein, incorporated in the 17-gene LSC score which predicts treatment response in AML[9] |
| *MN1* | 0.502 | (1.1E-23) | 0.565 | (<E-45) | A transcriptional coactivator, overexpression could induce AML in mice and predict ATRA resistance in human AML patients[43,44] |
| *ABCG1* | 0.504 | (6.7E-24) | 0.610 | (<E-45) | Belongs to ATP-binding cassette (ABC) superfamily; responsible for important chemoresistance mechanism in AML[45-50] |
| *ABCA2* | 0.367 | (1.5E-12) | 0.507 | (2.3E-11) | Belongs to ATP-binding cassette (ABC) superfamily; a strong prognostic biomarker for multidrug resistance in pediatric acute lymphoblastic leukemia[45-50] |
| *ABCB1* | 0.353 | (1.2E-11) | 0.364 | (5.2E-6) | Belongs to ATP-binding cassette (ABC) superfamily; responsible for important chemoresistance mechanism in AML[45-50] |
| *ABCC1* | 0.310 | (3.2E-9) | 0.458 | (5.2E-9) | Belongs to ATP-binding cassette (ABC) superfamily; responsible for important chemoresistance mechanism in AML[45-50] |

## Discussion

AML cells have abnormal genetic background, either mutations or aberrant expression of specific genes. In recent years, several gene expression scores have been proposed for prognostic prediction of AML patients. We previously developed a 11-gene mRNA expression signature, including *AIF1L, CXCR7*, DATT, *GPR56, H1F0, IFITM3, KIAA0125, MX1, STAB1, TM4SF1* and *TNS3*, for prognostication in AML patients[11]. Another group built a six-gene leukemia stem cell (LSC) score with the incorporation of *DNMT3B, GPR56, CD34, SOCS2, SPINK2*, and *KIAA0125* expressions for pediatric AML[32]. Recently, Ng et al. proposed a 17-gene LSC score that incorporated expressions of 17 stemness-related genes, including *KIAA0125*, and showed the scoring system was powerful to predict prognosis in AML patients[9]. Among these prognostic-relevant genes, *KIAA0125* is the only non-coding gene and expressed only in the homo sapiens, but not in mice. *KIAA0125* is located on chromosome 14 of the human genome. It was reported to be upregulated in amelobalstoma but shown as a tumor suppressor gene in colorectal cancer[51, 52]. However, its role in tumorigenesis is still largely unknown. In this study, we found that the expression level of *KIAA0125* in BM was significantly higher in AML patients than normal HSC transplant donors. The expression of *KIAA0125* was lower in the mature subtype M3, but higher in more immature subtypes, M0 and M1, compatible with the finding that it is an LSC-related gene[20]. The similar finding could be seen in the GSE 12662 cohort. Further bioinformatics study also showed highly significant association of *KIAA0125* expression with stem cell signatures, either HSC or LSC. We found that expressions of *SPINK2, MAP7, HOPX, MMRN1, DNMT3B, TCF4, SLC38A1, DOCK1, ARHGAP22, MN1*, and 4 genes in the ATP-binding cassette (ABC) superfamily *[ABCG1, ABCA2, ABCB1*, and *ABCC1)*, which have been reported to be associated with poor prognosis or chemoresistance in AML, were positively correlated to higher expression of *KIAA0125* (Figure 3b and Table 3). *HOPX, DOCK1, DNMT3B, MMRN1*, and *ARHGAP22* genes were reported as important leukemia stem cell markers[9, 34, 35, 37, 42]. Higher *SPINK2* expression was associated with poor prognosis in adult and pediatric AML[31, 32]. *TCF4* expression could predict outcome in *RUNX1*-mutated and translocated AML[39, 40]. *MN1* overexpression could induce AML in mice and predict ATRA resistance in human AML patients[43, 44]. Current knowledge about the association between theses *KIAA0125*-correlated genes and AML is summarized in Table 3. Interestingly, the expression levels of several ABC transporter genes, including *ABCA2*, *ABCB1, ABCC1, and ABCG1*, were also significantly higher in AML patients with higher *KIAA0125* expression. The ABC transporter family consists of 48 proteins in subfamilies designated A to G and some of them are known to be associated with multidrug resistance via ATP-dependent drug efflux[45, 46, 49]. *ABCB1, ABCC1* and *ABCG1* were reported to be responsible for chemoresistance in AML[45, 48]. The translational expression of *ABCA2* was shown to be a prognostic marker for drug resistance in pediatric acute lymphoblastic leukemia[47, 50]. The underlying mechanistic basis of the high correlation of these 4 genes to the expression of *KIAA0125* warrants further studies.

To the best of our knowledge, this is by far the first study specifically addressing the expression of lncRNA *KIAA0125* and its clinical and biological associations in AML patients. We found that higher *KIAA0125* expression were closely associated with *RUNX1* and *DNMT3A1* mutations in both the NTUH and TCGA cohorts. Patients with higher *KIAA0125* expression were more refractory to chemotherapy with a lower CR rate and higher refractory rate (Table 1). They had shorter OS and DFS not only among total cohort, but also in subgroups of patients with non-APL and those with normal karyotype. Based on its important clinical significance, *KIAA0125* could be a potential therapeutic target for AML and might give some directives to future work. Further experimental studies are necessary to delineate how *KIAA0125* participates in the stem cell biology of hematopoietic lineages and its role in the pathogenesis in AML.

## Data Availability

The raw and normalized microarray data reported in this article have been deposited in the Gene Expression Omnibus database (accession number GSE68469 and GSE71014).

https://www.ncbi.nlm.nih.gov/geo/query/acc.cgi?acc=GSE68469

https://www.ncbi.nlm.nih.gov/geo/query/acc.cgi?acc=GSE71014

## Acknowledgements

We would like to acknowledge the service provided by Department of Laboratory Medicine, Department of Medical Research, and Division of Hematology, Department of Internal Medicine, National Taiwan University Hospital.

## Authorship Contributions

YHW and CCL contribute equally to this study. YHW and CCL were responsible for data collection and management, statistical analysis and interpretation, literature research, and manuscript writing; SYH and CYY were responsible for data management and statistical analysis; CLH assisted in statistical analysis; SHL, CHT and HAH was responsible for data collection and management; and WCC and HFT planned, designed, and coordinated the study over the entire period and wrote the manuscript.

